# Arterial hypertension and β-amyloid accumulation have spatially overlapping effects on posterior white matter hyperintensity volume: A cross-sectional study

**DOI:** 10.1101/2022.07.19.22277546

**Authors:** Jose Bernal, Stefanie Schreiber, Inga Menze, Anna Ostendorf, Malte Pfister, Jonas Geisendörfer, Aditya Nemali, Anne Maass, Renat Yakupov, Oliver Peters, Lukas Preis, Luisa Schneider, Ana Lucia Herrera, Josef Priller, Eike Jakob Spruth, Slawek Altenstein, Anja Schneider, Klaus Fliessbach, Jens Wiltfang, Björn H. Schott, Ayda Rostamzadeh, Wenzel Glanz, Katharina Buerger, Daniel Janowitz, Michael Ewers, Robert Perneczky, Boris-Stephan Rauchmann, Stefan Teipel, Ingo Kilimann, Christoph Laske, Matthias H. Munk, Annika Spottke, Nina Roy, Laura Dobisch, Peter Dechent, Klaus Scheffler, Stefan Hetzer, Steffen Wolfsgruber, Luca Kleineidam, Matthias Schmid, Moritz Berger, Frank Jessen, Miranka Wirth, Emrah Düzel, Gabriel Ziegler

**Author notes:** Shared first authorship (Jose Bernal, Stefanie Schreiber). Shared last authorship (Emrah Düzel, Gabriel Ziegler). Correspondence to: Jose Bernal & Miranka Wirth, Full address: German Centre for Neurodegenerative Diseases (DZNE), Magdeburg, Leipziger Str. 44, 39120 Magdeburg, Germany & German Centre for Neurodegenerative Diseases (DZNE), Dresden, Tatzberg 41, 01307 Dresden, Germany, &.

## Abstract

**Background:** Posterior white matter hyperintensities (WMH) in subjects across the Alzheimer’s disease (AD) spectrum with minimal vascular pathology suggests that amyloid pathology—not just arterial hypertension—impacts WMH, adversely influencing cognition. Here we seek to determine the effect of both hypertension and Aβ positivity on WMH, and their impact on cognition.

**Methods:** We analysed data from subjects with a low vascular profile and normal cognition (NC), subjective cognitive decline (SCD), and amnestic mild cognitive impairment (MCI) enrolled in the ongoing observational multicentre DZNE Longitudinal Cognitive Impairment and Dementia Study (n=375, median age 70.2 [IQR 66.0-74.4] years; 176 female; NC/SCD/MCI 127/162/86). All subjects underwent a rich neuropsychological assessment. We focused on baseline memory and executive function—derived from multiple neuropsychological tests using confirmatory factor analysis—, baseline preclinical Alzheimer’s cognitive composite 5 (PACC5) scores, and changes in PACC5 scores over course of three years (ΔPACC5).

**Results:** Subjects with hypertension or Aβ positivity presented the largest WMH volumes (*p*_*FDR*_<0.05), with spatial overlap in the frontal (hypertension: 0.42±0.17; Aβ: 0.46±0.18), occipital (hypertension: 0.50±0.16; Aβ: 0.50±0.16), parietal lobes (hypertension: 0.57±0.18; Aβ: 0.56±0.20), corona radiata (hypertension: 0.45±0.17; Aβ: 0.40±0.13), optic radiation (hypertension: 0.39±0.18; Aβ: 0.74±0.19), and splenium of the corpus callosum (hypertension: 0.36±0.12; Aβ: 0.28±0.12). Hypertension, Aβ positivity, and WMH were connected to cognition. First, WMH coincided with worse cognitive performance and outcomes (*p*_*FDR*_<0.05), regardless of Aβ and hypertension. Accelerated cognitive decline was associated with WMH in the genu of the corpus callosum and segments of the forceps major and inferior fronto-occipital longitudinal fasciculus (*p*_*FDR*_<0.05). Second, hypertension was indirectly linked to cognitive performance at baseline and over time via splenial WMH (*indirect-only effect*; memory: −0.05±0.02, *p*_*FDR*_=0.029; executive: −0.04±0.02, *p*_*FDR*_=0.067; PACC5: −0.05±0.02, *p*_*FDR*_=0.030; ΔPACC5: −0.09±0.03, *p*_*FDR*_=0.043). Third, the relationship between Aβ positivity and baseline and longitudinal cognitive performance was independent of WMH burden.

**Conclusions:** Posterior white matter is susceptible to hypertension and Aβ accumulation and it mediates the association between hypertension and cognitive dysfunction. Posterior WMH could be a promising target to tackle the downstream damage related to the potentially interacting and potentiating effects of the two pathologies.

**Trial Registration:** German Clinical Trials Register (DRKS00007966, 04/05/2015)

## Background

The term “cerebral white matter hyperintensities” (WMH) describes dynamic and diffuse microstructural alterations in both periventricular and deep white matter, which appear hypodense on computed tomography and hyperintense on T2-weighted magnetic resonance imaging (MRI) and coincide with demyelination, axon loss, and gliosis [1,2]. WMH are common—especially but not exclusively in old age—and relate to a large spectrum of clinical symptoms, including apathy, fatigue, delirium, depression, progressive cognitive impairment, physical function disturbances, and increased risk of dementia and stroke [2,3].

Alterations to the functioning of cerebral micro-vessels—also known as cerebral small vessel disease (CSVD)—caused, for instance, by long-term exposure to cardiovascular risk factors (hypertension particularly) have been assumed to drive WMH formation [4–6]. Nonetheless, recent studies demonstrating elevated global and posterior WMH in patients along the Alzheimer’s disease (AD) spectrum with minimal vascular pathology (for review see [1,7–9]) call into question the assumption that any “AD-related” WMH solely reflect a vascular contribution. This viewpoint has also been challenged by studies reporting a more “AD-like” WMH pattern in Aβ positive subjective cognitive decline (SCD), mild cognitive impairment (MCI) or AD. The “AD-like” pattern is roughly confined to deep and periventricular posterior regions comprising the (parieto-)occipital lobe, corona radiata, optic (thalamic) radiation, or the corpus callosum (especially splenium) and presents neuropathologically with underlying gliosis, and axonal and myelin loss, but minimal vascular pathology—likely occurring secondary to cortical neurodegeneration [7–10]. Further studies demonstrate frontal or temporoparietal WMH dominance in AD as well and others report mixed results on the relationship between posterior WMH and AD pathology [11–14]. “WMH of presumed vascular origin” are, however, usually depicted in deep and periventricular frontal areas, suggesting some spatial WMH heterogeneity in hypertension compared to AD [8,15].

Here we use region- and voxel-based lesion analysis to determine the effect of both hypertension and AD pathology, i.e. β-amyloid (Aβ) positivity, on WMH as well as their interacting impact on cognition. For that purpose, we study WMH of non-demented participants of a large multicentre cohort with available cerebrospinal fluid (CSF) AD biomarkers, history of hypertension, and cross-sectional as well as longitudinal neuropsychological tests.

## Methods

### Study design

We used baseline MRI, CSF AD biomarkers, cognitive performance scores, medical records and longitudinal cognitive performance scores from the DELCODE (DZNE Longitudinal Cognitive Impairment and Dementia Study) cohort, an observational multicentre study from the German Centre for Neurodegenerative Diseases (DZNE) that focuses on the multimodal assessment of preclinical and clinical AD stages [16]. All participants received an extensive assessment at the local study site prior to joining DELCODE, which included medical history, psychiatric and neurological examination, neuropsychological testing, blood laboratory work-up, and routine MRI in accordance with local standards. All memory clinics used the Consortium to Establish a Registry for Alzheimer’s Disease (CERAD) neuropsychological test battery [17] to assess cognitive function. We focused on non-complaining healthy controls with normal cognition (NC) and participants with SCD and MCI and excluded patients with dementia due to AD to enrich our sample by variance due to vascular disease and Aβ pathology.

The presence of SCD and amnestic MCI was diagnosed using the existing research criteria for SCD [18,19] and MCI [20], respectively. Participants were diagnosed with SCD if they reported subjective cognitive decline or memory concerns, as expressed to the physician of the memory centre, and had a test performance better than −1.5 standard deviations (SD) below the age, sex, and education-adjusted normal performance on all subtests of the CERAD battery. The MCI group consisted of participants with amnestic MCI, as defined by age, sex, and education-adjusted performance below −1.5 SD on the delayed recall trial of the CERAD word-list episodic memory tests.

The NC group was recruited through local newspaper advertisements. Individuals who responded to the advertisement were screened by telephone with regard to SCD. The control group had to achieve unimpaired cognitive performance according to the same definition as the SCD group.

All participants entered DELCODE based on either their clinical diagnosis derived from the clinical workup or their identification as a control subject according to the procedures outlined. Additional inclusion criteria for all groups were age[≥[60 years, fluent German language skills, capacity to provide informed consent, and presence of a study partner. The main exclusion criteria for all groups were conditions clearly interfering with participation in the study or the study procedures, including significant sensory impairment. The following medical conditions were considered exclusion criteria: current major depressive episode, major psychiatric disorders either at baseline or in the past (e.g., psychotic disorder, bipolar disorder, substance abuse), neurodegenerative disorder other than AD, vascular dementia, history of stroke with residual clinical symptoms, history of malignant disease, severe or unstable medical conditions, and clinically significant laboratory abnormalities in vitamin B12. Prohibited drugs included chronic use of psychoactive compounds with sedative or anticholinergic effects, use of anti-dementia agents in SCD, amnestic MCI, and control subjects, and investigational drugs for the treatment of dementia or cognitive impairment one month before entry and throughout the duration of the study.

All participants gave written informed consent before inclusion in the study. DELCODE is retrospectively registered at the German Clinical Trials Register (DRKS00007966, 04/05/2015) and was approved by ethical committees and local review boards.

### Hypertension

Medical records were retrospectively screened for cardiovascular risk factors at the time of MRI. Patients diagnosed before to have primary or secondary arterial hypertension were considered to suffer from arterial hypertension (1: hypertensive; 0: normotensive). Single blood pressure measurements were not taken into account since repeated, long-term or at-home measurements would be required for the final diagnosis [21].

### Cognitive performance

All participants underwent a rich neuropsychological assessment, comprising the Mini-Mental State Examination (MMSE), Alzheimer’s Disease Assessment Scale–Cognitive 13-item subscale (ADAS-Cog 13), the Free and Cued Selective Reminding Test (FCRST; including a serial subtraction task, Wechsler Memory Scale revised version (WMS-R), Logical Memory [Story A] and Digit Span), two semantic fluency tasks (animals and groceries), the Boston Naming Test (15-item short version analogue to the CERAD battery, supplemented by five infrequent items from the long version), the oral form of the Symbol-Digit-Modalities Test (including a subsequent free recall of symbols and symbol-digit pairings), Trail Making Test Parts A and B, Clock Drawing and Clock Copying, a recall task of previously copied figures (as in the CERAD test battery), the Face Name Associative Recognition Test, and a Flanker task to assess executive control of attention. We focused on memory and executive function at baseline derived from these neuropsychological tests using confirmatory factor analysis to reduce the influence of test-specific effects and measurement errors [22].

We also leveraged the Preclinical Alzheimer’s Disease Cognitive Composite (PACC5) [23], which provides a single outcome measure reflective of episodic memory, timed executive function, and global cognition; domains that have been found sensitive to amyloid pathology. The PACC5 score was calculated as the mean of an individual’s z-standardised performance in the FCSRT Free Recall and Total Recall, the MMSE, the WMS-R Logical Memory Story A Delayed Recall, the Symbol-Digit-Modalities Test, and the sum of the two category fluency tasks and used the baseline mean and SD values of the cognitively unimpaired group of our sample to derive the subtest z-scores.

We selected subjects with available PACC5 scores over three annual follow-ups for further analysis. We estimated rates of change in these PACC5 scores over time using a linear mixed effect model (ΔPACC5 from hereon). We expressed it as follows:

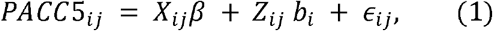

where *PACC*5_*ij*_ is the PACC5 scores of subject *i* ∈ [1, *N*] at visit *j* ∈ [1, *t*]; *X*_*ij*_ ∈ ℝ^*N*×*p*^ a matrix of the *p* predictor variables; *β* ∈ ℝ^*p*^ a vector of fixed-effects regression coefficients; *Z*_*ij*_ ∈ ℝ^*N*×*q*^ a design matrix for the *q* random effects; *b*_*ij*_∈ ℝ^*q*^ a vector of random effects; and *ϵ*_*ij*_ the within-subject measurement errors. The fixed effects structure include clinical group structure measured at baseline and their corresponding interaction with time (*t*_*ij*_).The fixed effects include age, sex and education taken at baseline.

### Structural MRI

Structural MRI scans were acquired at nine German DZNE sites on Siemens MR scanners (including three TIM Trio, four Verio, one Skyra, and one Prisma system). We used T1-weighted MPRAGE images (3D GRAPPA PAT 2, 1 mm^3^ isotropic, 256 × 256, 192 sagittal slices, repetition time 2500 ms, echo time 4.33 ms, inversion time 1100 ms, flip angle 7°, ∼5 min acquisition time) and T2-weighted 3D FLAIR images (GRAPPA PAT factor 2, 1 mm^3^ isotropic, 256 × 256, 192 sagittal slices, repetition time 5000 ms, echo time 394 ms, inversion time 1800 ms, ∼7 min acquisition time). Standard operating procedures, quality assurance and assessment were provided and supervised by the DZNE imaging network (iNET, Magdeburg) as described in [16]. We computed the mean background intensity as a surrogate measure of image quality and motion artefacts [24,25] and adjusted statistical models for it, as the quality of the scans determine also segmentation performance [26–28].

### Biomarker characterisation

Trained study assistants carried out lumbar punctures for 49% of all DELCODE participants. CSF samples were centrifuged, aliquoted and stored at −80°C for retests. Biomarkers known to mirror AD pathology (CSF Aβ42 and Aβ40) were determined by commercially available kits (V-PLEX Aβ Peptide Panel 1 (6E10) Kit (K15200E)). Each participant was classified as normal (-) or abnormal (+) with regard to amyloid levels based on the Aβ42/40 ratio, independently of their phosphorylated Tau (pTau) status, in line with the ATN classification system. Cut-offs (Aβ negative: Aβ42/40 >0.08; Aβ positive: Aβ42/40 ≤ 0.08) were calculated from DELCODE using the Gaussian mixture modelling in the R-package flexmix (v2.3-15) (for details see [16,29]).

### WMH segmentation and spatial processing

We processed baseline T1-weighted and FLAIR scans as follows. We performed bias field inhomogeneity correction, skull stripping, and segmentation using the Multi-Brain (MB) toolbox in statistical parametric mapping (SPM) [30]. We segmented grey matter (GM), white matter (WM), and CSF from T1-weighted scans with MB and identified WMH probability maps from FLAIR scans using the Lesion Prediction Algorithm in the Lesion Segmentation Toolbox [31]. We then used MB for normalising tissue classes (and WMH maps) to a DELCODE-specific MB template. We adjusted for local volume changes introduced by the normalisation in GM and WMH probability maps by modulation with Jacobian determinants [30,32]. Finally, we smoothed WMH maps with Gaussian kernels (6 mm full width at half maximum). Processing results of all steps were carefully checked visually and statistically using covariance-based tools provided in Computational Anatomy Toolbox 12 (CAT12) [33].

### ROI-based processing

We extracted WMH volume from 12 regions of interest (ROI) in cerebral WM, as described in detail in a previous study [11]. In brief, we created ROIs in accordance with the STRIVE criteria [34] and included the four lobes of the brain, four major WM tracks and three sections of the corpus callosum and a global cerebral WM mask. We calculated WMH volumes for each ROI and adjusted for total intracranial volume (TICV). All computations were conducted in the template space.

A schematic overview both processing and analysis methods is illustrated in **Figure S1**.

### Statistical analyses

#### Relationship between hypertension and Aβ positivity

We tested for associations between hypertension and Aβ positivity given their potential collinearity [35–38] using the Pearson’s Chi-squared test with Yates’ continuity correction in the R-package *stats* (v3.6.2).

#### Effects of hypertension and Aβ positivity on WMH

We hypothesised that a history of hypertension and an abnormal build-up of Aβ relate positively to the volume of WMH, but that both conditions display distinct spatial effects: hypertension on deep and periventricular frontal regions and Aβ on deep and periventricular posterior regions, as discussed in the literature [1,4–9]. We used a 2×2 ANCOVA model in CAT12 to examine the relationship between WMH segmentation maps (outcome) and hypertension and Aβ (factors) at a voxel level. Similarly, to probe the same relationship at an ROI level, we built 2×2 ANCOVA models in R (stats, v3.6.2), one for each region of interest separately. We controlled for covariates and confounders (see ‘*Covariates, confounders, and data transformation*’ below).

#### Effects of WMH on cognitive performance

Our hypothesis was that cognitive performance declined and rates of change in cognition increased as voxel-wise and regional WMH increased, in agreement with previous findings [2,3]. For voxel-based analysis, we used multiple linear regression in CAT12 with WMH segmentation maps as the dependent variable and cognitive performance as the independent variable. Likewise, for each region of interest, we also used multiple linear regression in R (stats, v3.6.2) to probe the relationship between regional WMH volume and cognition. We created different models with memory, executive function, PACC5, and ΔPACC5 as dependent variables. Note that, for studying the effect of baseline WMH on change in cognition, we leveraged summary statistics (ΔPACC5) instead of using a linear mixed effect model to keep the mass univariate analysis efficient [39] and both the voxel- and region-wise analyses consistent. We controlled for hypertension and Aβ positivity in addition to covariates and confounders (see ‘*Covariates, confounders, and data transformation’* below).

#### Mediation models

Assuming that long-term exposure to hypertension and Aβ build-up has a negative effect on the integrity of the white matter and that its damage—depicted in the form of regional WMH—impacts cognition negatively, we hypothesise that there is an indirect effect of hypertension and Aβ on cognition that is mediated by WMH volume, in line with theoretical considerations [5,6,40,41] (**Figure 1**). We used the R-package lavaan (v0.6-11) and followed the steps for mediation analysis suggested by Hair *et al*. [42] Significance was assessed using 95% confidence intervals generated by bias-corrected bootstrap with 1000 replicates.

**Figure 1.**
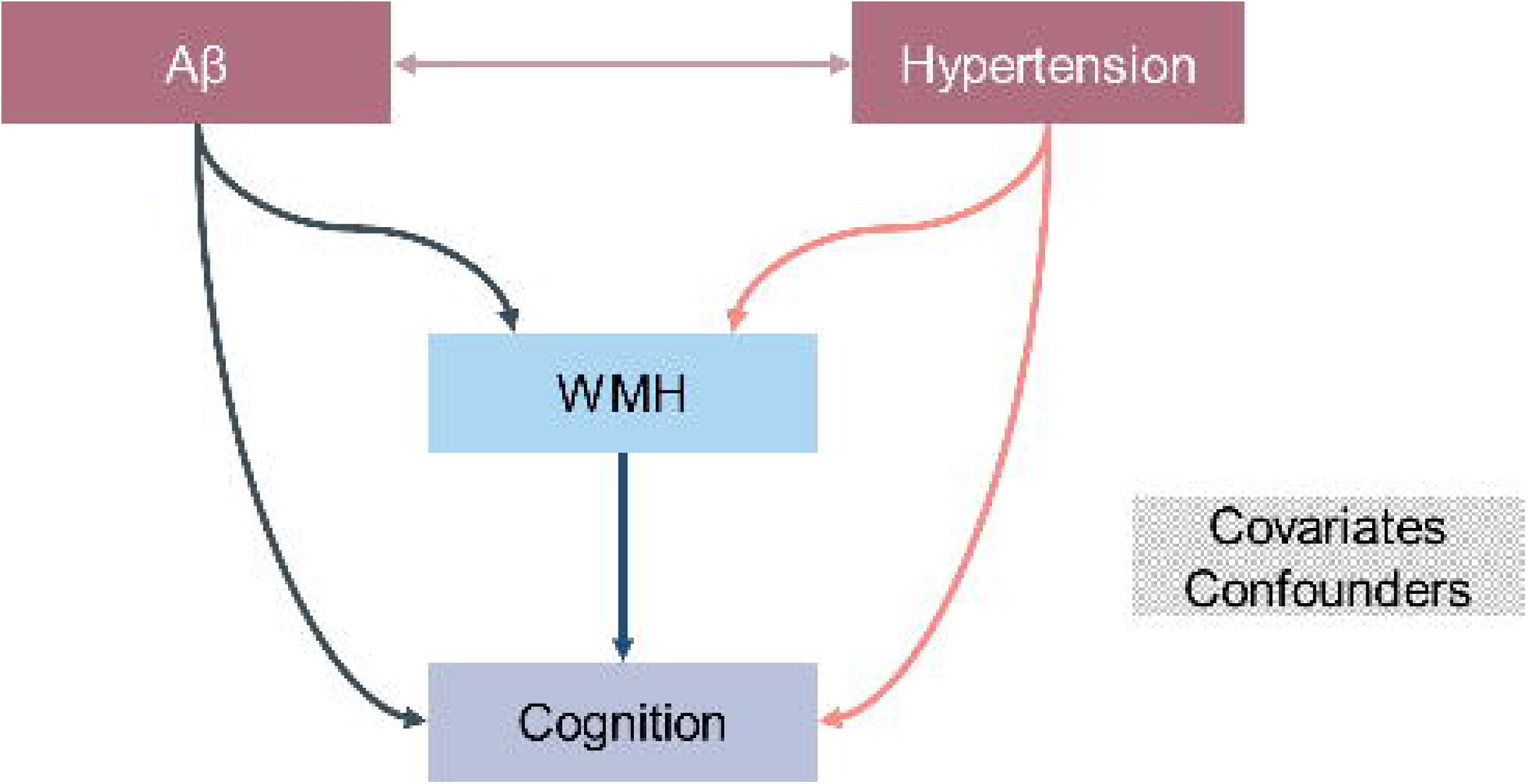
Model investigating direct and indirect (via WMH) effects of hypertension and Aβ positivity on cognition. Here we seek to understand whether subjects with arterial hypertension or Aβ positive status have worse cognitive performance at baseline (baseline memory, executive function, and PACC5 scores) and outcomes over time (ΔPACC5). Because both the Aβ and vascular pathologies may exacerbate the formation of WMH and these, in turn, may also contribute to brain dysfunction and poor cognitive outcomes [5,6,40,41], we also test for an indirect mediating effect of hypertension and Aβ positivity on cognitive performance via regional WMH volumes. We adjusted such models for age, sex, education, mean background intensity, and TICV, as described in ‘Covariates, confounders, and data transformation’.

#### Covariates, confounders, and data transformation

We adjusted all models for covariates (age, sex, years of education) and confounders (TICV) and mean background intensity to reduce biases brought in by correlated regressors. To account for collinearity between TICV and sex, we chose “overall mean” as “centring” for TICV and leveraged global scaling for this confounder. We refrained from adjusting our analyses for clinical groups to avoid collinearity issues with Aβ positivity (namely, Aβ positivity was more frequent in MCI vs NC and SCD). We log-transformed regional WMH volumes to account for skewness.

#### Explicit mask

We used an explicit mask to constrain the analysis to voxels in which data for at least five patients were available.

#### Correction for multiple comparisons

We adjusted *p*-values for multiple comparisons using the false discovery rates (FDR) approach to deal with the problem of multiple comparisons.[43]

## Results

### Sample description

We included baseline data of 375 subjects out of 1079 recruited for DELCODE after quality control and assessing the availability of CSF biomarkers and MRI (Figure S2; median age 70.0 [IQR 66.0-74.0] years, 46.9% female, median years of education 13 [IQR 12-17]; European origins). ΔPACC5 was only available for a subset (n=226/375). Demographics and global WMH volumes stratified by hypertension and Aβ positivity are summarised in **Table 1**. We found no significant association between arterial hypertension and Aβ positivity (Χ*2*=2.1302, *p*=0.1444).

**Table 1.** Demographics and WMH volume, stratified by hypertension diagnosis and Aβ positivity (n=375)^a^.

| Group | Subjects<br>n (%) | Age in years<br>Median [IQR] | Female<br>n (%) | Education in years<br>Median [IQR] | WMH volume in ml<sup>b</sup><br>Median [IQR] |
| --- | --- | --- | --- | --- | --- |
| Hypertension | 202 (53.9) | 72 [67, 75] | 89 (23.7) | 13 [12, 17] | 1.87 [0.83, 5.83] |
| Normotension | 173 (46.1) | 69 [64, 72] | 87 (23.2) | 14 [13, 17] | 1.19 [0.45, 2.78] |
| Aβ positive | 132 (35.2) | 72 [68, 76] | 55 (14.7) | 13 [12, 17] | 2.39 [1.11, 6.32] |
| Aβ negative | 243 (64.8) | 68 [65, 73] | 121 (32.2) | 13 [13, 17] | 1.16 [0.55, 2.96] |

<sup>a</sup>Global WMH volumes are higher subjects with hypertension vs normotension (W=13744, $p<0.001$ ) and Aβ positive vs negative (W=11851, $p<0.001$ ); age differences between groups explain, to a certain extent, these outcomes (hypertension vs normotension: W=13014, $p<0.001$ ; Aβ positive vs Aβ negative: W=10186, $p<0.001$ )
<sup>b</sup>Unadjusted WMH volumes in ml
n sample size
IQR interquartile range

### WMH are associated with both arterial hypertension and Aβ positivity

We initially investigated WMH in relation to hypertension and Aβ positivity. We observed that the global volume of WMH was a fourth greater in subjects with either a history of hypertension vs normotension (26% [95%-CI 5%, 52%]) or a positive vs negative Aβ status (25% [95%-CI 3%, 52%]) (**Table 2**). Regional variations in the frontal, parietal, and occipital—not temporal—lobes contributed to this outcome; regression coefficients for both hypertension and Aβ positivity were comparable in these three regions (**Table 2**). In posterior regions of the brain, we found that the relationship between WMH and hypertension was clearer than that between WMH and Aβ positivity in the splenium of the corpus callosum (**Figure 2** and **Table 2**); the opposite was true in the optic radiation (**Figure 2** and **Table 2**; peak: between forceps major and inferior fronto-occipital fasciculus).

**Table 2.**
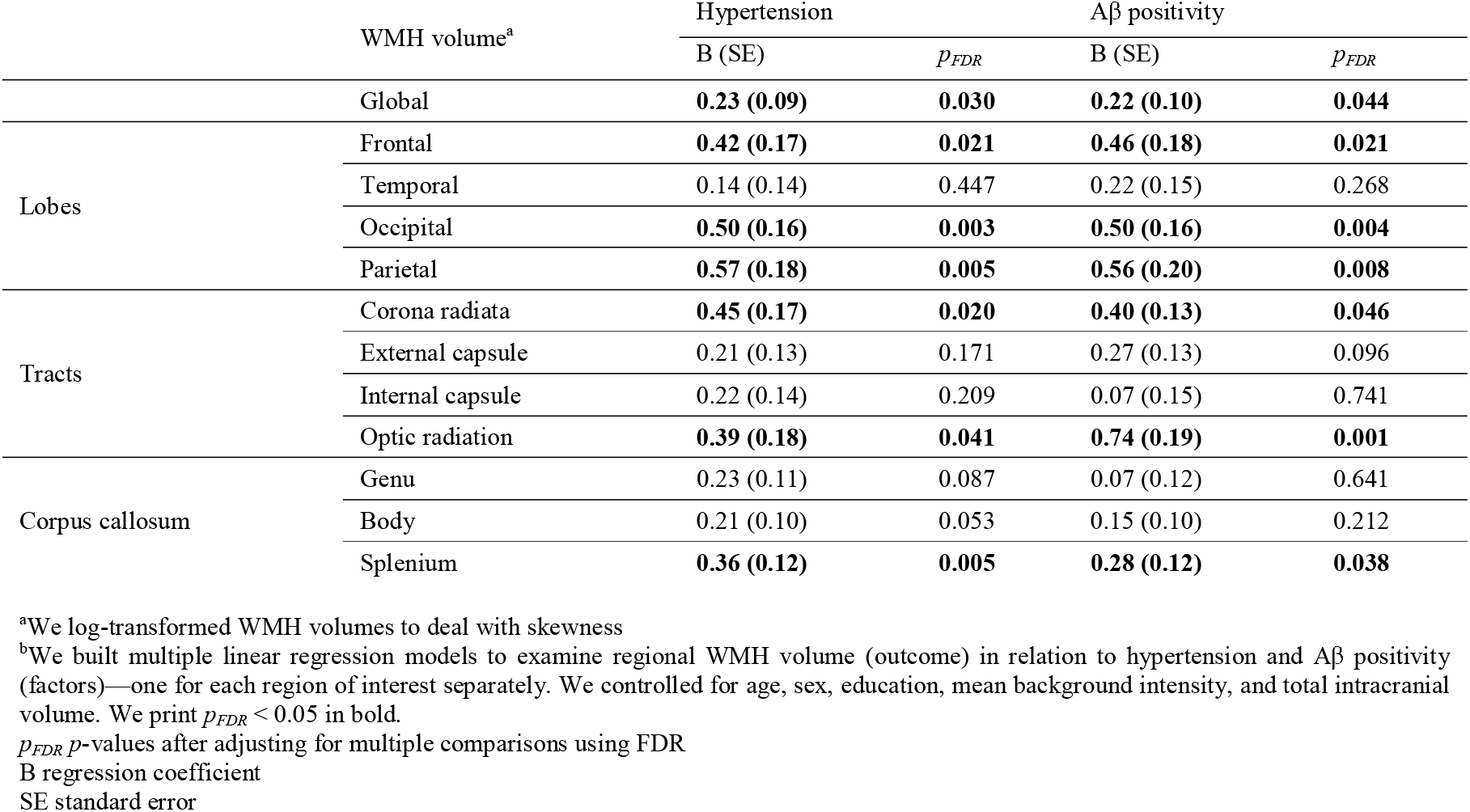
Subjects with hypertension and Aβ positivity present the largest frontal, parietal, and occipital WMH volumes^b^.

**Figure 2.**
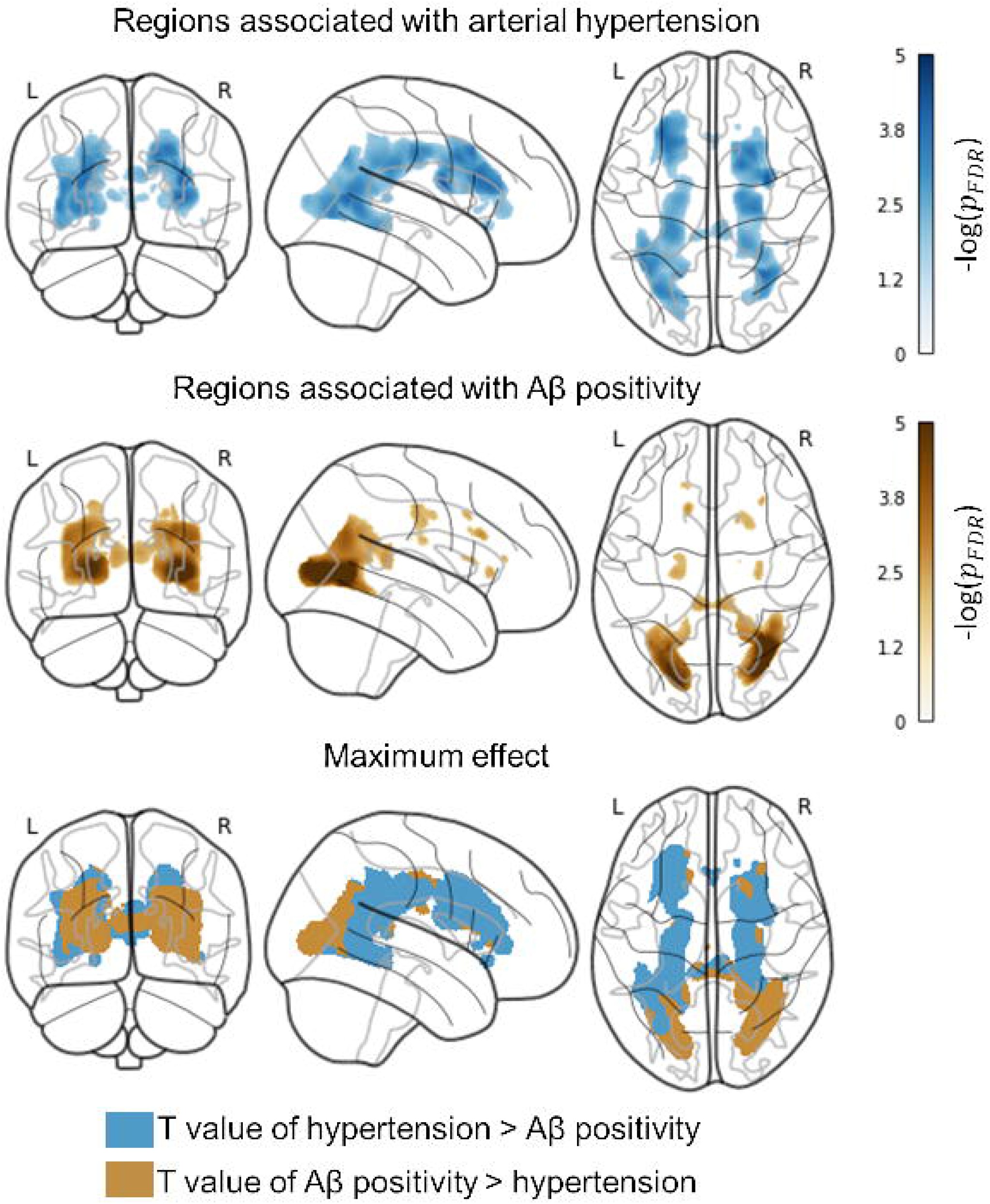
Posterior WMH probability is associated with both history of arterial hypertension and Aβ positivity. *Analysis*. We examined the relationship between WMH segmentation maps (outcome) and arterial hypertension and Aβ positivity (factors) at a voxel level via 2×2 ANCOVA. We accounted for the effects of age, sex, education, mean background intensity, and total intracranial volume. We used an explicit mask to constrain the analysis to voxels in which data for at least five patients were available. *Illustration*. Glass brain projections display regions where we found evidence for a link between WMH probability and hypertension and Aβ positivity (top and middle rows, respectively). In the bottom row, we coloured regions blue if *T* values for hypertension were greater than for Aβ positivity and gold otherwise. We thresholded contrast maps at 5% and adjusted *p*-values for FDR. *Findings*. Subjects with hypertension had significantly greater WMH volumes throughout the whole brain than those with normotension (peak: superior longitudinal fasciculus, *xyz*_*MNI*_ = [32, −1, 18], *T* = 3.88, *DoF* = [1.0, 367.0], *p*_*FDR*_ = 0.015). Moreover, WMH volume was significantly higher in subjects Aβ positivity versus negativity in posterior regions of the brain, particularly in segments of the forceps major and inferior fronto-occipital fasciculus (*xyz*_*MNI*_ = [30, −58, 4], *T* = 5.20, *DoF* = [1.0, 367.0], *p*_*FDR*_ = 0.001).

### WMH are negatively associated with cognitive performance and outcomes

We then investigated whether cognitive performance and outcomes were associated with WMH (**Figure 3** and **Table 3**). We found a significant association between global WMH volumes and worse baseline cognitive performance and a sharper cognitive decline over the course of three years, regardless of hypertension diagnosis and Aβ positivity (**Table 3**). Evidence for such a connection was present in most regions of interest, except in the external capsule. Such relationships were consistently evident around portions of the anterior thalamic radiation neighbouring the thalamus (**Figure 3**). In frontal and occipital regions, we also saw a significant link between WMH and quicker cognitive deterioration (**Figure 3 -** frontal peak at the level of the genu of the corpus callosum; occipital peak at the level of the forceps major and inferior fronto-occipital longitudinal fasciculus).

**Table 3.**
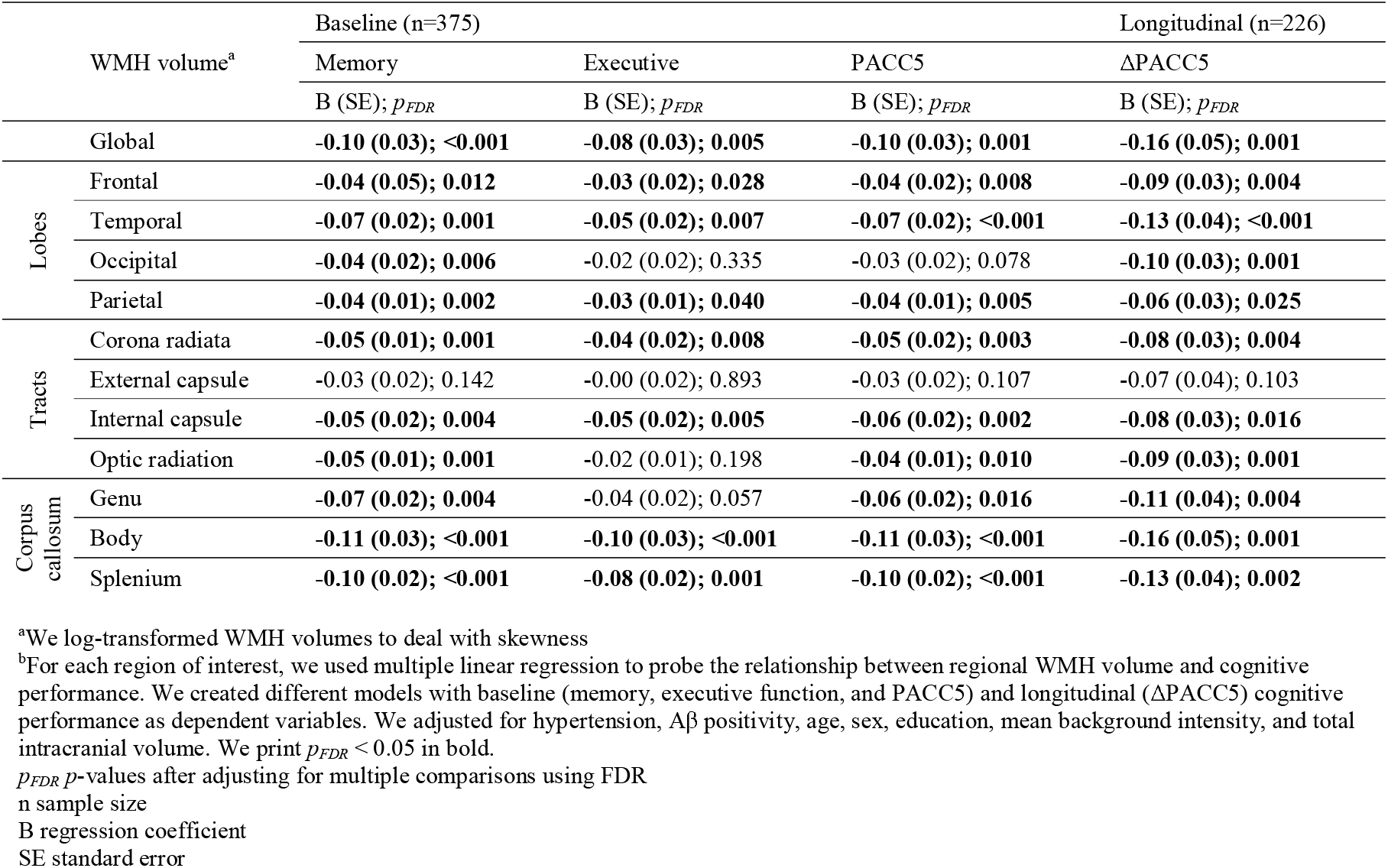
Higher WMH volumes are associated with worse and worsening cognitive performance^b^.

**Figure 3.**
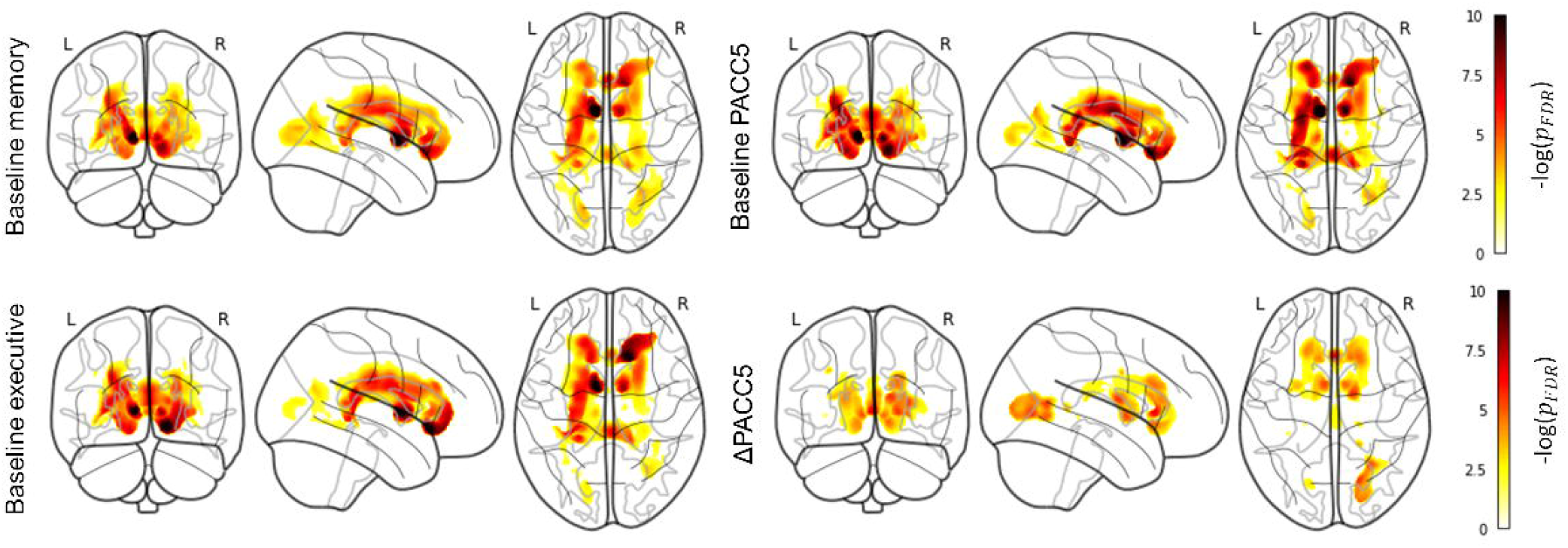
WMH volume is associated with worse baseline cognitive performance and accelarated decline over time. *Analysis*. We used multiple linear regression with WMH segmentation maps as the dependent variable and cognitive performance as the independent variable. We accounted for the effects of hypertension, Aβ positivity, age, sex, education, mean background intensity, and total intracranial volume. We used an explicit mask to constrain the analysis to voxels in which data for at least five patients were available. We thresholded contrast maps at 5% and adjusted *p*-values for FDR. *Illustration*. Regression results with memory (top left), executive function (bottom left), PACC5 (top right), and ΔPACC5 (bottom right) as independent variables. *Findings*. We found WMH to be significantly associated with worse cognitive performance at baseline and sharper decline within a three-year period. Such relationships were consistently evident around portions of the anterior thalamic radiation neighbouring the thalamus (memory: *xyz*_*MNI*_ = [−8, −1, 3], *T* = 7.00, *DoF* = [1.0, 366.0], *p*_*FDR*_ = 1.44×10^−5^; executive: *xyz*_*MNI*_ = [−9, 0, 5], *T* = 6.74, *DoF* = [1.0, 366.0], *p*_*FDR*_ = 2.85×10^−5^; PACC5: *xyz*_*MNI*_ = [−8, 1, 4], *T* = 7.20, *DoF* = [1.0, 366.0], *p*_*FDR*_ = 8.43×10^−6^; ΔPACC5: *xyz*_*MNI*_ = [−7, 2, 2], *T* = 4.53, *DoF* = [1.0, 217.0], *p*_*FDR*_ = 5.12×10^−3^). Frontal and occipital WMH also coincided with a faster cognitive decline (frontal peak at the level of the genu of the corpus callosum: *xyz*_*MNI*_ = [−1, 23, 4], *T* = 5.19, *DoF* = [1.0, 217.0], *p*_*FDR*_ = 1.37×10^−3^; occipital peak at the level of the forceps major and inferior fronto-occipital longitudinal fasciculus: *xyz*_*MNI*_ = [17, −81, 2], *T* = 4.69, *DoF* = [1.0, 217.0], *p*_*FDR*_ = 1.23×10^−2^).

### The effect of Aβ positivity on cognition does not depend on WMH, but that of hypertesion does

Our final assessment consisted of determining whether Aβ positivity or hypertension were associated with cognition (**Table 4**). Despite the lack of evidence for a direct association between hypertension and cognitive performance, we found hypertension to be indirectly linked to both worse performance at baseline and a steeper deterioration over the course of three years via splenial WMH (regression coefficient ± standard error; memory: −0.05±0.02, *p*_*FDR*_=0.029; executive: −0.04±0.02, *p*_*FDR*_=0.067; PACC5: −0.05±0.02, *p*_*FDR*_=0.030; ΔPACC5: −0.09±0.03, *p*_*FDR*_=0.043). On the other hand, the association between Aβ positivity and baseline and longitudinal cognitive performance (memory: −0.33±0.08, *p*_*FDR*_<0.001; executive: −0.21±0.08, *p*_*FDR*_<0.001; PACC5: −0.29±0.09, *p*_*FDR*_=0.006; ΔPACC5: −0.34±0.04, *p*_*FDR*_<0.05) was independent of its association with regional WMH volumes (no grounds for mediation in general, as shown in **Table 2**).

## Discussion

Using data from a large multi-site cohort of older adults along the AD spectrum (n=375), we investigated the impact of arterial hypertension and Aβ positivity on WMH and cognition. Our data suggest that (i) both hypertension and Aβ positivity are associated with increased volume of WMH at both voxel and regional levels, (ii) WMH are strongly associated with poor cognitive performance and outcomes, (iii) posterior WMH have a role in the association between hypertension and cognitive performance at baseline and over time, and (iv) the relationship between Aβ positivity and cognition does not depend on WMH. We observed a posterior WMH dominance in Aβ-positive older adults in the predementia stage of the AD continuum. Global and posterior WMH presence and volume were nonetheless the largest when both Aβ retention and hypertension occurred simultaneously and the smallest when none of them did. The posterior white matter could therefore be considered vulnerable to the independent yet interacting and potentiating effects of AD pathology and hypertension-related CSVD. One could thus consider posterior WMH to be a structural correlate that underlies the common observations that vascular disease, in particular hypertension, lowers the threshold for all-cause dementia development in face of pre-existing AD pathology, and vice versa [4–6]. As posterior WMH dominance could also relate to cerebrovascular deposition of Aβ, i.e. cerebral amyloid angiopathy (CAA), a condition that highly overlaps with AD pathology (for review see [44,45]), we visually inspected susceptibility-weighted sequences of all MRIs. Isolated lobar haemorrhagic markers were found in less than 10% (of them 19 were diagnosed with possible and 4 with probable CAA according to the Boston criteria [46,47]) of participants, making a relevant impact of CAA on posterior WMH in our sample highly unlikely.

WMH can negatively impact cognitive function, but associations with memory have been less consistent compared to associations with executive function (for review see [48]). With the exception of the external capsule, we found rather substantial evidence for associations between WMH and worse cognitive performance, likewise affecting memory and executive function, and outcomes (**Table 3**). Intriguingly, hypertension was associated with executive, memory, and baseline and longitudinal global cognitive function only via splenial WMH, a white matter structure responsible for cognitive processing and a hub where distinct pathologies impact the neural circuitries interconnecting the temporal and occipital regions of both cerebral hemispheres [7,49–51]. White matter damage in this region, as associated with cardiovascular risk, could be expected to translate to lower cognitive functioning in global cognition but also in discrete domains [7]. In previous studies though, posterior/splenial WMH have been found associated with executive (including attention), but not memory function [7,10]. Differences may arise from WMH quantification methods and/or smaller sample sizes including AD patients only (not individuals with SCD/MCI), in whom largely advanced (medial temporal lobe) AD pathology is the major driver for memory decline, possibly “diluting” concurrent memory effects of posterior WMH.

Contrary to our expectations and to strong evidence from large longitudinal population-based studies (for review see [6]), we did not see a direct effect of hypertension effect on cognition but rather an indirect-only effect via splenial WMH. This finding might reflect a selection bias of the DELCODE study: exclusion of individuals with advanced vascular disease, which would likewise result in the exclusion of those with severe and uncontrolled hypertension. This constellation, additionally, explains the somewhat lower prevalence of arterial hypertension (nearly 54% compared to 63%), with a slightly higher number of Aβ positives (35% compared to a range of 17% to 34%) compared to that in population-based cohorts aged over 60 years [52–54]. Our definition of arterial hypertension was based on retrospective screening of medical records for already existing hypertension diagnoses, which might have missed those participants with recently, i.e. newly, diagnosed hypertension after baseline MRI, also contributing to lower prevalence.

This study has limitations. First, our imaging results are cross-sectional. While our findings suggest WMH are indeed spatially associated with both hypertension and Aβ positivity, they do not address causality (e.g. vascular risk first, impaired brain drainage second). Longitudinal analysis of DELCODE imaging data might provide further insights into the influence of lifestyle over time and help disentangle the mixed effects observed in this cross-sectional study. Second, our mediation model investigates whether WMH volume can mediate the association between Aβ and hypertension on cognitive function. While this choice was based on a theoretical consideration [5,6,40,41], a model where the AD and CSVD pathologies (here as Aβ accumulation and WMH burden) cyclically contribute to each other would also be feasible [2,5,55]. Third, the study of WMH probability patterns in other cohorts of individuals (e.g. whose origins are other than European; DELCODE participants are predominantly of European origins) with a high vascular but low AD profile or vice versa could be informative on the mechanisms leading to these findings in a more general way. Further, we did not consider WMH patterns, which could be punctuated or confluent, for example, or the clinically established distinction between deep and periventricular WMH.

## Conclusion

Our work points toward a large spatial overlap between the effect of arterial hypertension and Aβ build-up on WMH, with both constellations considered risk factors for white matter damage. At the same time, our work calls into question whether posterior WMH are a core feature related to AD pathology, alternatively suggesting that posterior white matter is vulnerable to both vascular and amyloid pathologies. While the effect of Aβ on cognition would seem rather independent of WMH, posterior WMH seem to play a role in the association between arterial hypertension and poor cognitive performance and outcomes; it could be a promising target to tackle the downstream damage related to the interacting and potentiating effect of multiple pathologies.

## Data Availability

All data produced in the present work are contained in the manuscript

## Abbreviations

Aβ: β-amyloid
AD: Alzheimer’s disease
ATN: Amyloid/Tau/Neurodegeneration
B: regression coefficient
CAA: Cerebral amyloid angiopathy
CERAD: Consortium to Establish a Registry for Alzheimer’s Disease
CN: Non-complaining healthy controls
CSF: Cerebrospinal fluid
CSVD: Cerebral small vessel disease
FDR: False discovery rate
FLAIR: Fluid Attenuated Inversion Recovery
GM: Grey matter
IQR: Interquartile range
MCI: Mild cognitive impairment
MRI: Magnetic resonance imaging
NC: Normal cognition
PACC5: Preclinical Alzheimer’s Cognitive Composite 5
ROI: Region of interest
SCD: Subjective cognitive decline
SD: Standard deviation
SE: Standard error
SPM: Statistical parametric mapping
TICV: Total intracranial volume
WM: White matter
WMH: White matter hyperintensities

## Declarations

### Consent for publication

Not applicable.

### Availability of data and materials

The datasets used and analysed during the current study are available from the corresponding author on reasonable request.

### Author’s contributions

Conceptualisation: JB, SS, MW, GZ. Methodology: JB, SS, MW, GZ. Software: JB, GZ. Formal analysis: JB. DELCODE study design: ED, AS, and FJ. Image processing: JB, GZ, RY, MW. Image analysis and modelling: JB, GZ. Investigation: JB, SS, MW, GZ. Writing original draft preparation: JB, SS, MW, GZ. Writing – review and editing: All authors.

## Acknowledgements

We would like to express our gratitude to all DELCODE participants. We also thank the Max-Delbrück-centrum für Molekulare medizin in der Helmholtz-Gemeinschaft (MDC), Freie Universität Berlin Center for Cognitive neuroscience Berlin (CCNB), Bernstein Center für Computional Neuroscience Berlin, Universitätsmedizin Göttingen Core Facility MR-Research Göttingen, Institut für Klinische Radiologie Klinikum der Universität München, and Universitätsklinikum Tübingen MR-Forschungszentrum.

