## Supplementary Material for "Arterial hypertension and β-amyloid accumulation have spatially overlapping effects on posterior white matter hyperintensity volume: A cross-sectional study"

**Figure S1. Schematic illustration of WMH processing and analysis pipeline.** We segmented WMH based on baseline T1w and FLAIR scans using the Lesion Prediction Algorithm in the Lesion Segmentation Toolbox. We then used Multi-Brain (MB) for normalising tissue classes (and WMH maps) into a DELCODE-based MB template. At this stage, we integrated WMH over 12 regions of interest, including the whole brain, lobes, corona radiata, external capsule, internal capsule, optic radiation, and corpus callosum. In R, we used these estimates to perform ROI-based analysis (regional WMH volume in relation to hypertension, Aβ positivity, and cognitive performance) and mediation models (hypertension and Aβ as independent variables; regional WMH volume as mediator; and cognitive performance as dependent variable). Later, we adjusted for local volume changes introduced by the normalisation in WMH probability maps via modulation. Finally, we smoothed WMH maps with Gaussian kernels (6 mm full width at half maximum). We used the modulated and smoothed WMH maps as input to our voxel-based WMH analysis. We investigated local effects of hypertension and Aβ on WMH maps and of WMH on cognition. We used CAT12 for this purpose.

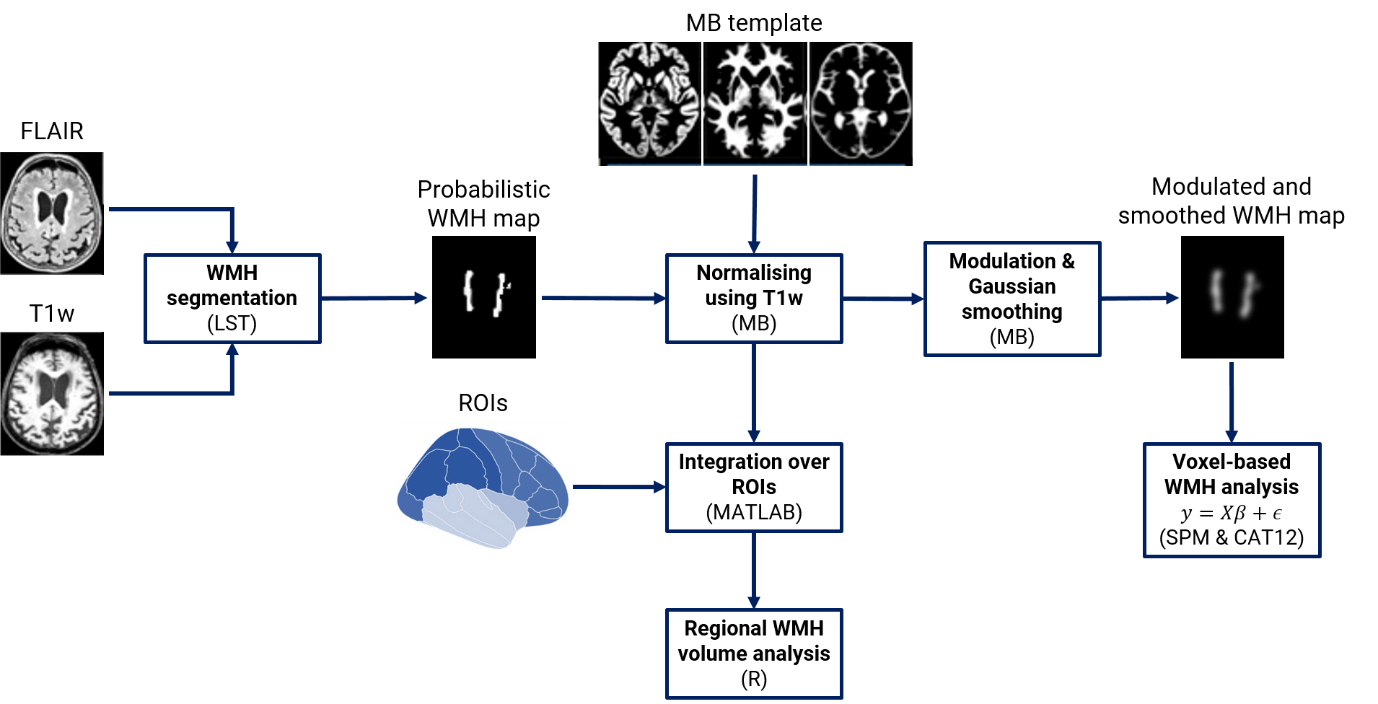

**Figure S2. Inclusion/exclusion flowchart.** We included non-AD subjects with cerebrospinal fluid (CSF) biomarkers, MRI data, and information regarding cardiovascular risk factors.

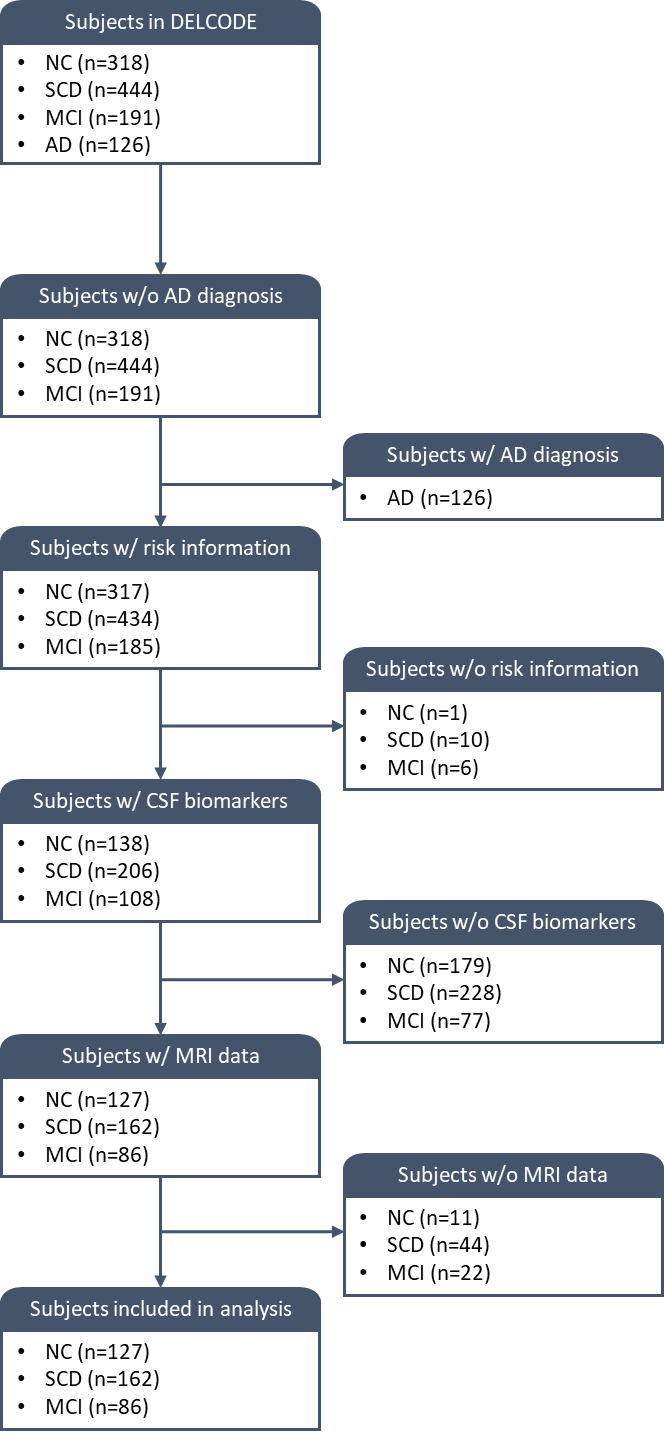
