## Supplementary material for "Arterial hypertension and β-amyloid accumulation have spatially overlapping effects on posterior white matter hyperintensity volume: A cross-sectional study": Table 4

**Table 4. Both hypertension and Aβ positivity are associated with worse cognitive performance at baseline and a sharper decline over the course of three years. However, while WMH mediated the connection between hypertension and cognition, they did not seem to play a role in that between Aβ positivity and cognition**^b^

| **Dependent variable** | **Mediator variable**^a^ | **Hypertension** | **Aβ positivity** | **Hypertension** | **Aβ positivity** | **Hypertension** | **Aβ positivity** |
| --- | --- | --- | --- | --- | --- | --- | --- |
|  |  | **Direct effect** | **Indirect effect** | **Direct effect** | **Indirect effect** | **Direct effect** | **Indirect effect** |
|  |  | **B (SE); *p_FDR_*** | **B (SE); *p_FDR_*** | **B (SE); *p_FDR_*** | **B (SE); *p_FDR_*** | **B (SE); *p_FDR_*** | **B (SE); *p_FDR_*** |
| Baseline memory (n=375) | Global | 0.06 (0.07); 0.433 | -0.03 (0.02); 0.083 | 0.03 (0.07); 0.666 | **-0.33 (0.07); <0.001** | -0.03 (0.02); 0.133 | **-0.36 (0.08); <0.001** |
|  | Frontal | 0.05 (0.07); 0.500 | -0.02 (0.01); 0.162 | 0.03 (0.07); 0.666 | **-0.33 (0.08); <0.001** | -0.02 (0.02); 0.179 | **-0.36 (0.08); <0.001** |
|  | Occipital | 0.07 (0.07); 0.433 | -0.04 (0.02); 0.083 | 0.03 (0.07); 0.666 | **-0.33 (0.08); <0.001** | -0.03 (0.02); 0.117 | **-0.36 (0.08); <0.001** |
|  | Parietal | 0.07 (0.07); 0.433 | -0.04 (0.02); 0.083 | 0.03 (0.07); 0.666 | **-0.33 (0.08); <0.001** | -0.03 (0.02); 0.111 | **-0.36 (0.08); <0.001** |
|  | Corona radiata | 0.06 (0.07); 0.452 | -0.03 (0.02); 0.106 | 0.03 (0.07); 0.666 | **-0.33 (0.08); <0.001** | -0.02 (0.02); 0.129 | **-0.36 (0.08); <0.001** |
|  | Optic radiation | 0.06 (0.07); 0.464 | -0.03 (0.02); 0.128 | 0.03 (0.07); 0.666 | **-0.31 (0.08); <0.001** | **-0.05 (0.02); 0.029** | **-0.36 (0.08); <0.001** |
|  | Splenium | 0.08 (0.07); 0.349 | **-0.05 (0.02); 0.029** | 0.03 (0.08); 0.666 | **-0.32 (0.08); <0.001** | -0.04 (0.02); 0.102 | **-0.36 (0.08); <0.001** |
| Baseline executive (n=375) | Global | -0.06 (0.07); 0.455 | -0.03 (0.01); 0.112 | -0.09 (0.07); 0.294 | **-0.21 (0.08); 0.029** | -0.02 (0.02); 0.209 | **-0.23 (0.08); 0.014** |
|  | Frontal | -0.07 (0.07); 0.433 | -0.02 (0.01); 0.208 | -0.09 (0.07); 0.294 | **-0.21 (0.08); 0.030** | -0.02 (0.02); 0.256 | **-0.23 (0.08); 0.014** |
|  | Occipital | -0.07 (0.07); 0.383 | -0.01 (0.01); 0.387 | -0.09 (0.07); 0.294 | **-0.22 (0.08); 0.023** | -0.01 (0.01); 0.433 | **-0.23 (0.08); 0.014** |
|  | Parietal | -0.06 (0.07); 0.451 | -0.02 (0.02); 0.180 | -0.09 (0.07); 0.294 | **-0.21 (0.08); 0.029** | -0.02 (0.01); 0.274 | **-0.23 (0.08); 0.014** |
|  | Corona radiata | -0.07 (0.07); 0.452 | -0.025 (0.01); 0.134 | -0.09 (0.07); 0.294 | **-0.21 (0.08); 0.029** | -0.02 (0.02); 0.190 | **-0.23 (0.08); 0.014** |
|  | Optic radiation | -0.08 (0.07); 0.370 | -0.01 (0.01); 0.370 | -0.09 (0.07); 0.294 | **-0.21 (0.08); 0.029** | -0.02 (0.02); 0.365 | **-0.23 (0.08); 0.014** |
|  | Splenium | -0.05 (0.07); 0.542 | -0.04 (0.02); 0.067 | -0.09 (0.07); 0.294 | **-0.20 (0.08); 0.035** | -0.03 (0.02); 0.180 | **-0.23 (0.08); 0.014** |
| asline PACC5 (n=375) | Global | 0.08 (0.07); 0.368 | -0.03 (0.02); 0.083 | 0.04 (0.07); 0.547 | **-0.29 (0.08); 0.006** | -0.03 (0.02); 0.149 | **-0.32 (0.09); <0.001** |
|  | Frontal | 0.07 (0.07); 0.405 | -0.03 (0.02); 0.151 | 0.05 (0.07); 0.547 | **-0.29 (0.09); 0.006** | -0.03 (0.02); 0.178 | **-0.32 (0.09); <0.001** |
|  | Occipital | 0.07 (0.07); 0.420 | -0.02 (0.01); 0.164 | 0.05 (0.07); 0.547 | **-0.29 (0.09); 0.006** | -0.02 (0.02); 0.215 | **-0.32 (0.09); <0.001** |
|  | Parietal | 0.07 (0.07); 0.379 | -0.03 (0.02); 0.102 | 0.05 (0.07); 0.547 | **-0.28 (0.09); 0.006** | -0.03 (0.02); 0.149 | **-0.32 (0.09); <0.001** |
|  | Corona radiata | 0.07 (0.07); 0.379 | -0.03 (0.02); 0.098 | 0.05 (0.07); 0.547 | **-0.29 (0.09); 0.006** | -0.03 (0.02); 0.128 | **-0.32 (0.09); <0.001** |
|  | Optic radiation | 0.07 (0.07); 0.432 | -0.02 (0.01); 0.155 | 0.05 (0.07); 0.547 | **-0.27 (0.09); 0.012** | -0.04 (0.02); 0.067 | **-0.32 (0.09); <0.001** |
|  | Splenium | 0.10 (0.07); 0.279 | **-0.05 (0.02); 0.030** | 0.05 (0.07); 0.547 | **-0.27 (0.09); 0.012** | -0.04 (0.02); 0.120 | **-0.32 (0.09); <0.001** |
| ΔPACC5 (n=226) | Global | -0.10 (0.13); 0.496 | -0.06 (0.03); 0.095 | -0.16 (0.13); 0.294 | **-0.35 (0.14); 0.041** | -0.04 (0.03); 0.370 | **-0.38 (0.15); 0.029** |
|  | Frontal | -0.11 (0.13); 0.448 | -0.05 (0.03); 0.172 | -0.16 (0.13); 0.294 | -0.33 (0.14); 0.058 | -0.06 (0.04); 0.208 | **-0.38 (0.15); 0.029** |
|  | Occipital | -0.09 (0.13); 0.547 | -0.08 (0.04); 0.084 | -0.16 (0.13); 0.294 | -0.32 (0.14); 0.069 | -0.06 (0.03); 0.134 | **-0.38 (0.15); 0.029** |
|  | Parietal | -0.10 (0.13); 0.486 | -0.06 (0.03); 0.121 | -0.16 (0.13); 0.294 | **-0.34 (0.14); 0.050** | -0.04 (0.03); 0.208 | **-0.38 (0.15); 0.029** |
|  | Corona radiata | -0.11 (0.13); 0.464 | -0.06 (0.03); 0.121 | -0.16 (0.13); 0.294 | **-0.34 (0.14); 0.043** | -0.03 (0.03); 0.352 | **-0.38 (0.15); 0.029** |
|  | Optic radiation | -0.10 (0.13); 0.471 | -0.06 (0.03); 0.162 | -0.16 (0.13); 0.294 | -0.30 (0.14); 0.083 | -0.07 (0.03); 0.083 | **-0.38 (0.15); 0.029** |
|  | Splenium | -0.08 (0.13); 0.572 | **-0.09 (0.03); 0.043** | -0.16 (0.13); 0.294 | **-0.34 (0.14); 0.048** | -0.04 (0.02); 0.250 | **-0.38 (0.15); 0.029** |

^a^We log-transformed WMH volumes to deal with skewness

^b^We tested for indirect mediating effects of arterial hypertension and Aβ positivity (independent variables) on cognitive performance (dependent variable) via regional WMH volume (mediator variable) (**Figure 1**). The significance of *p*-values for these associations was based on 95% confidence intervals generated using bias-corrected bootstrap with 1000 replicates. We controlled for hypertension, age, sex, education, mean background intensity, and total intracranial volume. We print *p_FDR_* < 0.05 in bold.

*p_FDR_* *p*-values after adjusting for multiple comparisons using FDR

n sample size

B regression coefficient

SE standard error
